# Postoperative complications of transcatheter and surgical aortic valve replacement in patients with borderline pulmonary hypertension:A retrospective cohort study

**DOI:** 10.64898/2025.12.14.25342232

**Authors:** Dan Zhao, Yu Wang, Lingyu Kuai, Xisheng Shan, Wei Zhang, Bo Liu, Xiaowen Meng, Ke Peng, Fuhai Ji, Hao Cheng, Yufan Yang

**Affiliations:** Department of Anesthesiology, First Affiliated Hospital of Soochow University, Suzhou, Jiangsu, People’s Republic of China; Institute of Anesthesiology, Soochow University, Suzhou, Jiangsu, People’s Republic of China; Otorhinolaryngology Department, China Academy of Chinese Medical Science Eye Hospital, Beijing, China; Department of Anesthesiology, The third affiliated hospital of Zhengzhou University, Henan, China

**Keywords:** Transcatheter aortic valve replacement, Surgical aortic valve replacement, Postoperative composite complications, Borderline pulmonary hypertension, Propensity-score matching analysis

## Abstract

**Background:** Borderline pulmonary hypertension (mean pulmonary artery pressure 20–24 mmHg) is increasingly recognized as a clinically significant condition that may influence perioperative outcomes in patients undergoing aortic valve replacement. This study aims to explore the 30-day composite complication rates and mortality among patients with borderline pulmonary hypertension who received transcatheter aortic valve replacement (TAVR) or surgical aortic valve replacement (SAVR).

**Methods:** The retrospective cohort study included 194 patients with borderline pulmonary hypertension who underwent either TAVR (n = 67) or SAVR (n = 127) between 2012 and 2023. Propensity score matching (1:1) was used to balance baseline characteristics. The primary outcome was a composite of 30-day postoperative complications and mortality. Secondary outcomes included 1-year mortality, individual complications, and perioperative clinical parameters.

**Results:** The 30-day composite complications and mortality were 56 of 67 (83.6%) patients undertaking TAVR, compared with 101 of 127 (79.5%) patients taking SAVR. After matching, 42 patients were included in each group. There was no significant difference in the incidence of the primary composite outcome between TAVR and SAVR groups (85.7% vs. 85.7%). One-year mortality was also comparable (2.4% vs. 0%). TAVR was associated with a higher rate of cardiovascular complications (21.4% vs. 4.8%) but a lower rate of renal complications (0% vs. 26.2%). TAVR patients had shorter operative time, earlier extubation, reduced blood loss, and shorter intensive care unit and hospital stays, but higher hospitalization costs.

**Conclusions:** In patients with borderline pulmonary hypertension, TAVR and SAVR demonstrated similar rates of 30-day composite complications and 1-year mortality. These findings support a support individualized treatment decisions to valve replacement strategy in this clinically vulnerable population.

## Introduction

Aortic valve disease remains a leading contributor to cardiovascular morbidity and mortality globally. Since the first performance of surgical aortic valve replacement (SAVR) in the 1960s, therapeutic strategies have advanced markedly with the advent of transcatheter aortic valve replacement (TAVR) [1, 2]. Initially, TAVR was restricted to patients at extreme or very high surgical risk; however, based on robust randomized clinical trials, its indications have expanded to encompass high-, intermediate-, and even low-risk populations [3–7]. Current clinical guidelines recommend a comprehensive decision-making framework that integrates clinical, anatomical, and surgical factors, eliminating strict contraindications for either procedure in most cases [5].

Among patients with aortic valve disease, borderline pulmonary hypertension, characterized by a mean pulmonary artery pressure of 20–24 mmHg, represents a critical yet underinvestigated hemodynamic phenotype [8]. It is associated with subclinical right ventricular remodeling and dysfunction, increased perioperative complications (e.g., right heart failure, respiratory failure), and worse long-term outcomes [8, 9]. For patients with aortic valve disease and borderline pulmonary hypertension, who are at the tipping point of progressive right heart failure, the choice between minimally invasive TAVR and open SAVR may have disproportionate implications for hemodynamic adaptation, organ function, and recovery [10–12].

Despite its clinical relevance, borderline pulmonary hypertension has been largely overlooked in comparative effectiveness research on TAVR vs. SAVR [13, 14]. SAVR requires cardiopulmonary bypass (CPB), which can exacerbate pulmonary vascular resistance and right ventricular dysfunction, key vulnerabilities in patients with borderline pulmonary hypertension, where the right heart-pulmonary vascular axis is already compromised [15]. TAVR’s minimally invasive approach may reduce perioperative stress, but it is linked to procedure-specific complications such as high-degree atrioventricular block and coronary obstruction [16, 17], which could disproportionately impact patients with borderline pulmonary hypertension-related cardiac reserve impairment. Additionally, the effect of both procedures on renal function, critical in borderline pulmonary hypertension due to the high prevalence of cardiorenal syndrome [9, 18], remains undefined.

Prior randomized clinical trials have established TAVR’s non-inferiority to SAVR in low-to-intermediate risk patients with aortic stenosis for outcomes including all-cause mortality or disabling stroke at 1–3 years [3–5], with durable benefits observed in low-risk cohorts [4]. However, these studies rarely stratify by borderline pulmonary hypertension status, leaving clinicians without evidence to guide treatment in this subgroup. To address this gap, this study analyzed a cohort of patients with borderline pulmonary hypertension undergoing TAVR or SAVR was analyzed, with their postoperative composite complications, mortality, and clinical outcomes compared to inform evidence-based, personalized care.

## Methods

### Study Design and Population

A retrospective cohort study was carried out involving patients with borderline pulmonary hypertension who received TAVR or SAVR at two branches of the First Affiliated Hospital of Soochow University from March 2012 to September 2023. The study design was reviewed and approved by the hospital’s ethics committee (No. 2025-993), and the trial was registered in the Chinese Clinical Trial Registry (available at http://www.chictr.org.cn, with the registration number ChiCTR 2500111590).

The inclusion criteria were patients over 50 years old undergoing TAVR or SAVR with borderline pulmonary hypertension. The key exclusion criteria included patients with other concomitant heart diseases or valvular diseases, synchronous receipt of additional cardiovascular surgery, left ventricular ejection fraction <20%, myocardial infarction or cerebral infarction within 1 month, liver and kidney dysfunction, preoperative sepsis, procedural failure or significant complications, such as paravalvular leak, valve thrombosis, or graft occlusion; concomitant malignant tumors; and data missing more than 20%. Among 194 enrolled patients, 67 (34.5%) were assigned to the TAVR cohort, and the remaining 127 (65.5%) to the SAVR cohort.

### Procedure

Under general anesthesia, a midline sternal incision was made to establish cardiopulmonary bypass (CPB) for SAVR. In the SAVR group, 4 mechanical heart valves and 8 bioprostheses were used, including the St. Jude Medical heart valve, St. Jude Medical Regent heart valve, Medtronic Open Pivot AP, Medtronic Mosaic and Edwards Perimount Magna Ease. Implanted devices included bileaflet mechanical valves and current-generation bioprostheses; sutures were placed in a sub-annular, pledget-reinforced fashion. Transthoracic echocardiography was used to measure the performance of the new valve perioperatively.

All procedures of TAVR were done in our hybrid operating room with a multidisciplinary team and a primed CPB circuit on a pump stand-by. The four different Chinese self-expanding brands of TAVR prostheses used were the Venus A-valve, TaurusOne, VitaFlow, and J-Valve. The surgery was performed under general anesthesia, and the best approach was selected based on preprocedural peripheral vascular assessment results. Transfemoral and transapical methods were mainly used in our center. Notably, the J-Valve was used exclusively via a transapical approach. With rapid cardiac pacing to lower systemic blood pressure, the valve was implanted after ensuring the correct position by transesophageal echocardiography (TEE) and fluoroscopy.

The data were accessed for research purposes in 10/11/2025. All baseline, perioperative, and follow-up information was collected by an independent investigator via the Lex Clinical Data Application 3.2 (Hangzhou Lejiu Healthcare Technology Co., Ltd.) within the Hospital Health Information System. Data at the 30-day and 1-year time points were gathered during in-clinic consultations or over the phone. These data were all entered into an electronic database and analyzed by a separate, independent biostatistician.

### Outcome Measures

The primary outcome was a composite endpoint of mortality and major complications (including infectious, respiratory, neurologic, cardiovascular, renal, thrombotic, gastrointestinal and surgical complications) within 30 days in patient with borderline pulmonary hypertension after TAVR or SAVR (Table S1). The secondary outcomes were defined as the one-year mortality and each component of the primary outcome within 30 days after TAVR or SAVR.

Intraoperative fluid volume, allogeneic blood transfusion volume, surgical time, postoperative valve function, extubation time, hospitalization expenses, and length of stay in the intensive care unit and surgical ward were reported as exploratory outcomes between the TAVR and SAVR groups. Demographics, complications, perioperative electrocardiogram, echocardiography, hemodynamics, laboratory, and medication data were also obtained and compared with the two groups.

### Sample size calculation

Age, gender, ASA classification, previous medical history, preoperative hemoglobin, preoperative albumin, C-reactive protein, operative duration, extubation time, length of stay in intensive care unit, and postoperative hospital stay are expected to be included as confounding factors in the model analysis.. According to previous literature report, the occurrence rate of major composite complications was 65% [19]. The calculation formula for sample size was 11*10/0.65, so the minimum sample size to be included was 170 patients.

### Statistical analysis

All data analyses were performed using R 4.1 software. Continuous variables following a normal distribution were expressed as mean ± standard deviation (SD), whereas those with non-normal distribution were summarized as median (interquartile range, IQR). For between-group comparisons of continuous data including demographic details and hospital-related characteristics, either Student’s t-test or the Mann-Whitney U test was applied, depending on data distribution. Categorical variables were displayed as absolute numbers or percentages, and intergroup differences were tested for statistical significance using Pearson’s chi-square test or Fisher’s exact test. Propensity score matching (PSM) was conducted based on logistic regression to balance baseline characteristics between the TAVR and SAVR groups, employing a 1:1 matching strategy with a caliper value of 0.25. Matching covariates included age, ASA classification, medical history, preoperative hemoglobin concentrations, and preoperative C-reactive protein concentrations. Logistic regression models were utilized to analyze the primary outcome measures. Kaplan-Meier (KM) curves were plotted to illustrate the 1-year overall survival rate of patients, and group differences in survival were compared via the log-rank test. Statistical significance was defined as a two-sided P-value < 0.05.

## Results

### Baseline characteristics of the patients

603 patients complying with the inclusion criteria were screened from the database between March 2012 and September 2023. Among them, 404 patients were eliminated according to exclusion criteria. The specific reasons for exclusion included 11 patients with concomitant congenital heart diseases, 237 with valvular diseases, 119 who were undergoing synchronous additional cardiovascular surgery, 9 with a left ventricular ejection fraction less than 20%, 3 with myocardial infarction, 1 with cerebral infarction within the past month, 1 with liver dysfunction, 8 with kidney dysfunction, 1 with preoperative sepsis, and 10 due to severe data loss. Additionally, 3 patients taking alternative surgery owing to valve displacement (n=2) and severe ventricular fibrillation (n=1), and one patient who had concurrent malignant gastric tumors (n=1). The analysis included 194 patients eventually (SAVR, n=127; TAVR, n=67). The complete flow chart of participant selection was presented in Fig 1. Table 1 outlines the baseline characteristics of all participants in the study. The majority of patient-related traits were comparable between the two groups, including age, ASA classification, coronary heart disease, hemoglobin, albumin, and C-reactive protein. Before PSM, the TAVR group had a significantly older age (72.3 ± 6.5 vs. 65.1 ± 7.8 years, P < 0.001), higher proportion of ASA III-IV classification (82.1% vs. 58.3%, P = 0.002), and lower preoperative hemoglobin (11.8 ± 1.5 vs. 12.6 ± 1.3 g/dL, P = 0.003) compared with the SAVR group. After PSM, all baseline variables were well-balanced (all P > 0.05, Table 1).

**Table 1.**
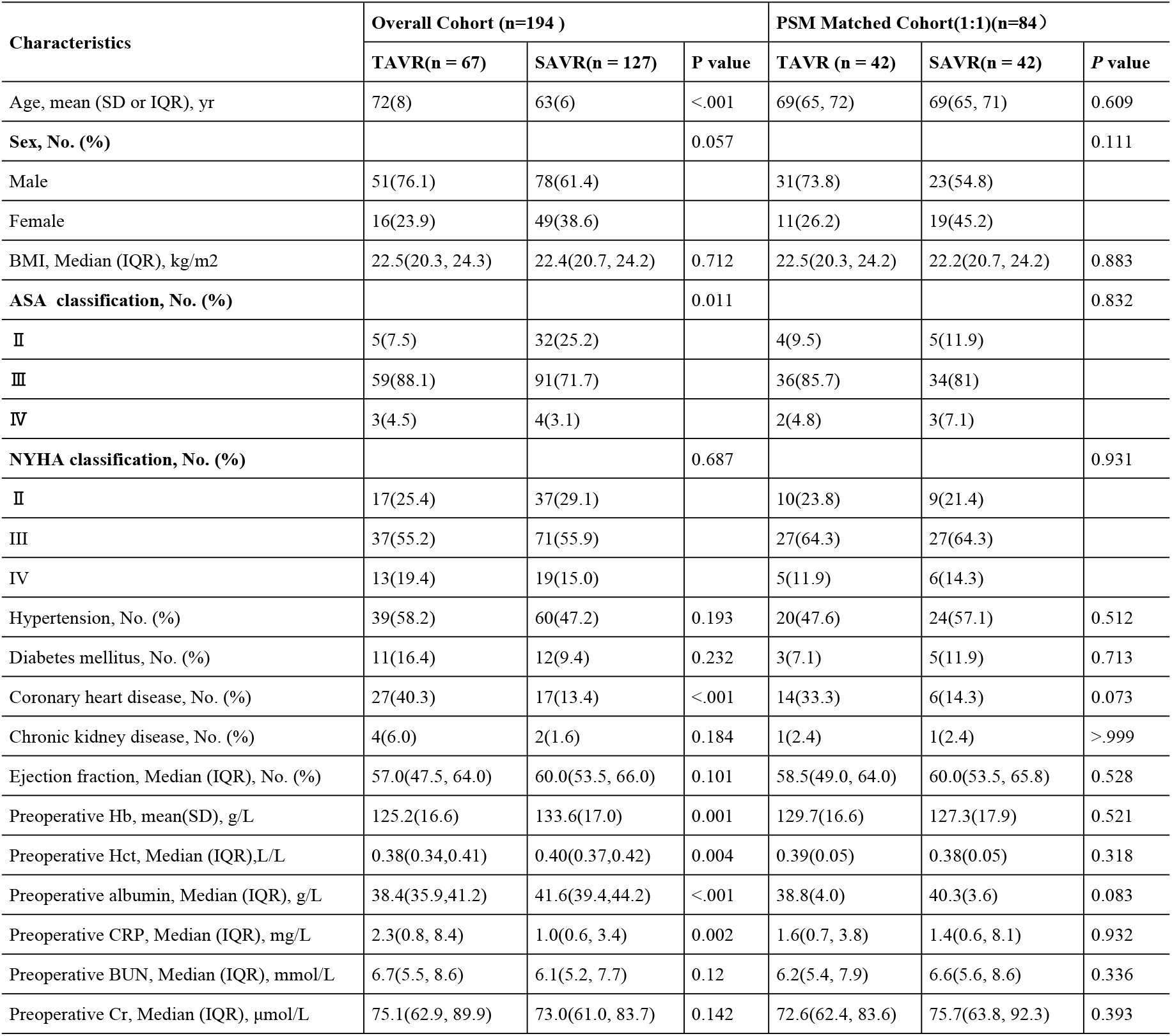
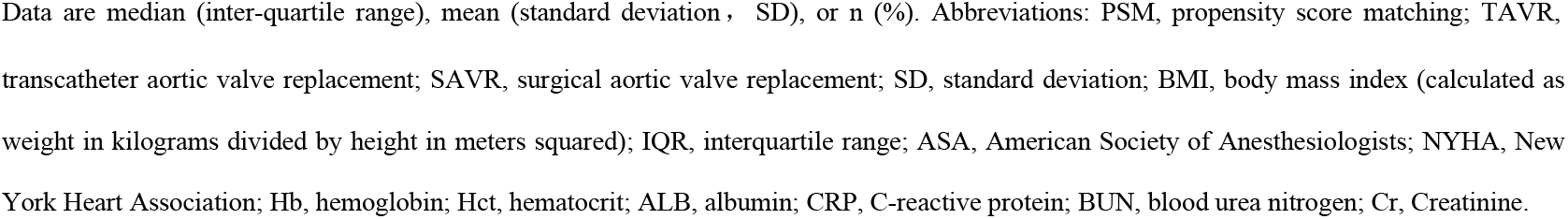
Baseline characteristics.

| Characteristics | Overall Cohort (n=194 ) |  |  | PSM Matched Cohort(1:1)(n=84) |  |  |
| --- | --- | --- | --- | --- | --- | --- |
|  | TAVR(n = 67) | SAVR(n = 127) | P value | TAVR (n = 42) | SAVR(n = 42) | P value |
| Age, mean (SD or IQR), yr | 72(8) | 63(6) | <.001 | 69(65, 72) | 69(65, 71) | 0.609 |
| **Sex, No. (%)** |  |  | 0.057 |  |  | 0.111 |
| Male | 51(76.1) | 78(61.4) |  | 31(73.8) | 23(54.8) |  |
| Female | 16(23.9) | 49(38.6) |  | 11(26.2) | 19(45.2) |  |
| BMI, Median (IQR), kg/m<sup>2</sup> | 22.5(20.3, 24.3) | 22.4(20.7, 24.2) | 0.712 | 22.5(20.3, 24.2) | 22.2(20.7, 24.2) | 0.883 |
| **ASA classification, No. (%)** |  |  | 0.011 |  |  | 0.832 |
| II | 5(7.5) | 32(25.2) |  | 4(9.5) | 5(11.9) |  |
| III | 59(88.1) | 91(71.7) |  | 36(85.7) | 34(81) |  |
| IV | 3(4.5) | 4(3.1) |  | 2(4.8) | 3(7.1) |  |
| **NYHA classification, No. (%)** |  |  | 0.687 |  |  | 0.931 |
| II | 17(25.4) | 37(29.1) |  | 10(23.8) | 9(21.4) |  |
| III | 37(55.2) | 71(55.9) |  | 27(64.3) | 27(64.3) |  |
| IV | 13(19.4) | 19(15.0) |  | 5(11.9) | 6(14.3) |  |
| Hypertension, No. (%) | 39(58.2) | 60(47.2) | 0.193 | 20(47.6) | 24(57.1) | 0.512 |
| Diabetes mellitus, No. (%) | 11(16.4) | 12(9.4) | 0.232 | 3(7.1) | 5(11.9) | 0.713 |
| Coronary heart disease, No. (%) | 27(40.3) | 17(13.4) | <.001 | 14(33.3) | 6(14.3) | 0.073 |
| Chronic kidney disease, No. (%) | 4(6.0) | 2(1.6) | 0.184 | 1(2.4) | 1(2.4) | >.999 |
| Ejection fraction, Median (IQR), No. (%) | 57.0(47.5, 64.0) | 60.0(53.5, 66.0) | 0.101 | 58.5(49.0, 64.0) | 60.0(53.5, 65.8) | 0.528 |
| Preoperative Hb, mean(SD), g/L | 125.2(16.6) | 133.6(17.0) | 0.001 | 129.7(16.6) | 127.3(17.9) | 0.521 |
| Preoperative Hct, Median (IQR),L/L | 0.38(0.34,0.41) | 0.40(0.37,0.42) | 0.004 | 0.39(0.05) | 0.38(0.05) | 0.318 |
| Preoperative albumin, Median (IQR), g/L | 38.4(35.9,41.2) | 41.6(39.4,44.2) | <.001 | 38.8(4.0) | 40.3(3.6) | 0.083 |
| Preoperative CRP, Median (IQR), mg/L | 2.3(0.8, 8.4) | 1.0(0.6, 3.4) | 0.002 | 1.6(0.7, 3.8) | 1.4(0.6, 8.1) | 0.932 |
| Preoperative BUN, Median (IQR), mmol/L | 6.7(5.5, 8.6) | 6.1(5.2, 7.7) | 0.12 | 6.2(5.4, 7.9) | 6.6(5.6, 8.6) | 0.336 |
| Preoperative Cr, Median (IQR), μmol/L | 75.1(62.9, 89.9) | 73.0(61.0, 83.7) | 0.142 | 72.6(62.4, 83.6) | 75.7(63.8, 92.3) | 0.393 |

Data are median (inter-quartile range), mean (standard deviation , SD), or n (%). Abbreviations: PSM, propensity score matching; TAVR, transcatheter aortic valve replacement; SAVR, surgical aortic valve replacement; SD, standard deviation; BMI, body mass index (calculated as weight in kilograms divided by height in meters squared); IQR, interquartile range; ASA, American Society of Anesthesiologists; NYHA, New York Heart Association; Hb, hemoglobin; Hct, hematocrit; ALB, albumin; CRP, C-reactive protein; BUN, blood urea nitrogen; Cr, Creatinine.

**Fig 1.**
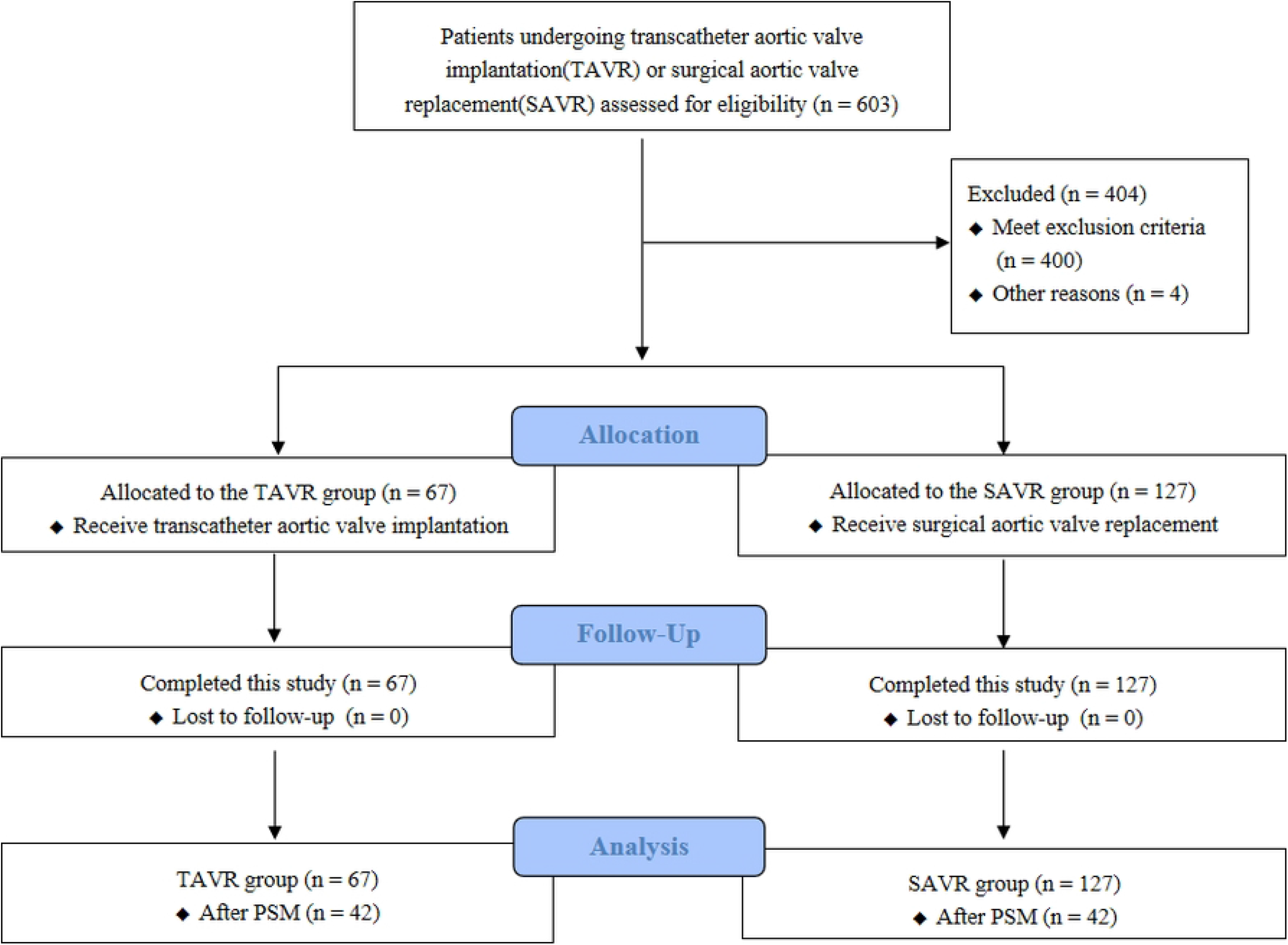
Flow diagram of study selection process.

### Primary outcome

The postoperative complication and mortality rate data were presented in Table 2. A primary composite complications within 30 days after surgery was confirmed for 56 of 67 (83.6%) patients undertaking TAVR, compared with 101 of 127 (79.5%) patients taking SAVR (P=0.623). After propensity score matching, the TAVR group retained 42 patients, while the SAVR group also retained 42 patients. Thirty-six patients (85.7%) in both groups experienced postoperative complications.

**Table 2.** Primary Outcome and Secondary Outcomes within 30 days and 1 year.

| Variable | Overall Cohort (n =194 ) |  |  | PSM Matched Cohort(1:1)(n=84) |  |  |
| --- | --- | --- | --- | --- | --- | --- |
|  | TAVR(n=67) | SAVR(n=127) | P value | TAVR(n=42) | SAVR(n= 42) | P value |
| **Primary Outcome** |  |  |  |  |  |  |
| Major composite complications, No. (%) | 56(83.6) | 101(79.5) | 0.623 | 36(85.7) | 36(85.7) | >.999 |
| **Secondary Outcomes** |  |  |  |  |  |  |
| Infectious complications, No. (%) | 22(32.8) | 43(33.9) | >.999 | 19(45.2) | 18(42.9) | >.999 |
| Respiratory complications, No. (%) | 41(61.2) | 82(64.6) | 0.759 | 25(59.5) | 30(71.4) | 0.359 |
| Neurologic complications, No. (%) | 4(6.0) | 7(5.5) | >.999 | 1(2.4) | 3(7.1) | 0.616 |
| Cardiovascular complications, No. (%) | 20(29.9) | 12(9.4) | 0.001 | 9(21.4) | 2(4.8) | 0.048 |
| Renal complications, No. (%) | 4(6.0) | 24(18.9) | 0.017 | 0 | 11(26.2) | <.001 |
| Thromboembolic complications, No. (%) | 0 (0) | 0 (0) | >.999 | 0 (0) | 0 (0) | >.999 |
| Gastrointestinal complications, No. (%) | 0 | 1(0.8) | >.999 | 0 (0) | 1(2.4) | >.999 |
| Surgical complications, No. (%) | 4(6.0) | 10(7.9) | 0.774 | 3(7.1) | 5(11.9) | 0.713 |
| 30-day mortality, No. (%) | 4(6.0) | 0 | 0.013 | 0 | 0 | >.999 |
| 1-year mortality, No. (%) | 5(7.5) | 3(2.4) | 0.127 | 1(2.4) | 0 | >.999 |

Date are n (%). PSM, propensity score matching; Abbreviations: TAVR, transcatheter aortic valve replacement; SAVR, surgical aortic valve replacement.

### Primary composite outcome under logistic regression

Based on the statistical results of the baseline indicators and clinical considerations, several factors including age, gender, ASA classification, previous medical history, preoperative hemoglobin levels, C-reactive protein levels, albumin levels, extubation time, operation duration, intensive care unit length of stay and postoperative hospital stay were incorporated into the logistic regression model (Table 3). Variables were included according to the standard of P<0.1. Univariate analysis revealed that the prematching variables included gender, serum albumin concentration, extubation time, intensive care unit length of stay, and postoperative hospital stay. The ASA classification met the standard after matching. The significant impact of age on surgical outcomes should also be considered [9]. The variables noted above were finally included in the multifactor logistic regression analysis (Table 4). Before and after propensity score matching, there was no remarkable variation in the primary composite outcome between the TAVR and SAVR groups. This indicates that clinical outcomes were analogous for borderline pulmonary hypertension patients receiving TAVR or SAVR.

**Table 3.** Univariate logistic regression analyses for the composite endpoint of major complications and deaths before and after propensity matching.

| Variables | Overall Cohort (n =194 ) |  | PSM Matched Cohort(1:1)(n=84) |  |
| --- | --- | --- | --- | --- |
|  | OR (95% CI) | P value | OR (95% CI) | P value |
| Exposure(TAVR) | 1.31 (0.62-2.95) | 0.495 | 1.20 (0.36-4.07) | 0.763 |
| Age, yr | 1.03 (0.98-1.08) | 0.259 | 1.01 (0.91-1.10) | 0.924 |
| Female | 0.52 (0.25-1.08) | 0.078 | 1.11 (0.33-3.97) | 0.872 |
| ASA III | 1.85 (0.77-4.23) | 0.152 | 6.40 (1.35-29.60) | 0.016 |
| ASA IV | 0.93 (0.17-7.19) | 0.933 | 1.60 (0.11-42.32) | 0.736 |
| Past medical history | 1.76 (0.85-3.64) | 0.127 | 1.90 (0.57-6.49) | 0.29 |
| Preoperative Hb, g/L | 1.00 (0.98-1.02) | 0.859 | 1.00 (0.96-1.03) | 0.782 |
| Preoperative CRP, mg/L | 1.05 (0.99-1.14) | 0.191 | 1.05 (0.97-1.21) | 0.424 |
| Preoperative ALB, g/L | 0.86 (0.78-0.94) | <.001 | 0.88 (0.72-1.04) | 0.144 |
| Extubation time, hour | 1.03 (1.01-1.05) | 0.016 | 1.02 (1.00-1.06) | 0.225 |
| Operative duration, min | 1.00 (0.99-1.01) | 0.258 | 1.00 (0.99-1.01) | 0.844 |
| Length of stay in ICU, hour | 1.01 (1.00-1.02) | 0.003 | 1.01 (1.00-1.03) | 0.023 |
| postoperative hospital stay, day | 1.07 (1.03-1.12) | 0.002 | 1.06 (1.01-1.14) | 0.062 |

Abbreviations: PSM, propensity score matching; OR, odds ratio; CI, confidence interval; TAVR, transcatheter aortic valve replacement; ASA, American Society of Anesthesiologists; Hb, hemoglobin; CRP, C-reactive protein; ALB, albumin; ICU, Intensive care unit.

**Table 4.** Multivarible logistic regression analyses for the composite endpoint of major complications and deaths before and after propensity matching.

| Variables | Overall Cohort (n =194 ) |  | PSM Matched Cohort(1:1)(n=84) |  |
| --- | --- | --- | --- | --- |
|  | OR (95% CI) | P value | OR (95% CI) | P value |
| Exposure(TAVR) | 1.42 (0.45-4.69) | 0.557 | 5.27 (0.89-40.24) | 0.080 |
| Age, yr | 0.98 (0.92-1.05) | 0.652 | 1.01 (0.88-1.14) | 0.939 |
| Female | 0.38 (0.15-0.92) | 0.033 | 1.76 (0.30-13.91) | 0.554 |
| ASA III | 1.50 (0.53-4.06) | 0.427 | 7.56 (1.06-60.55) | 0.044 |
| ASA IV | 0.62 (0.07-6.80) | 0.671 | 0.62 (0.01-29.46) | 0.792 |
| Preoperative Hb, g/L | 0.80 (0.69-0.91) | 0.001 | 0.93 (0.74-1.16) | 0.524 |
| Extubation time, hour | 1.01 (0.99-1.05) | 0.322 | 0.99 (0.95-1.06) | 0.816 |
| Length of stay in ICU, hour | 1.01 (1.00-1.02) | 0.062 | 1.02 (1.00-1.04) | 0.052 |
| Postoperative hospital stay, day | 1.07 (1.02-1.13) | 0.011 | 1.06 (0.99-1.16) | 0.165 |

Abbreviations: PSM, propensity score matching; OR, odds ratio; CI, confidence interval; TAVR, transcatheter aortic valve replacement; ASA, American Society of Anesthesiologists; Hb, hemoglobin; ICU, Intensive care unit.

### Secondary and other outcomes

In the TAVR group, 20 patients (29.9%) experienced cardiovascular complications, while in the SAVR group, 12 patients (9.4%) experienced cardiovascular complications (P=0.001). Among renal-related complications, the incidence was 6.0% in the TAVR group and 18.9% in the SAVR group (P=0.017). After matching, 9 patients (21.4%) experienced cardiovascular complications in TAVR group, and 2 patients (4.8%) experienced cardiovascular complications in SAVR group (P=0.048). There were no renal complications in the TAVR group, while in the SAVR group, there were 11 cases (26.2%) (P<0.001) after matching. Statistically significant differences did not happen in other related complications after matching. Regarding patient mortality at 30 days after surgery, there were 4 patients (6.0%) in the TAVR group 30 days after surgery, while there were no patients in the SAVR group (P=0.013). One year after surgery, 5 patients (7.5%) in TAVR group died, and 3 patients (2.4%) in SAVR group did (P=0.623). K−M curves estimating the one-year survival of the two groups were presented in Fig 2. The data after matching indicated that there were no deaths in either group 30 days after surgery, and only one patient in TAVR group died one year after surgery. The comparisons of 30-d and 1-year mortality revealed no significant differences in patients with borderline pulmonary hypertension undergoing TAVR versus SAVR. (Table 2).

**Fig 2.**
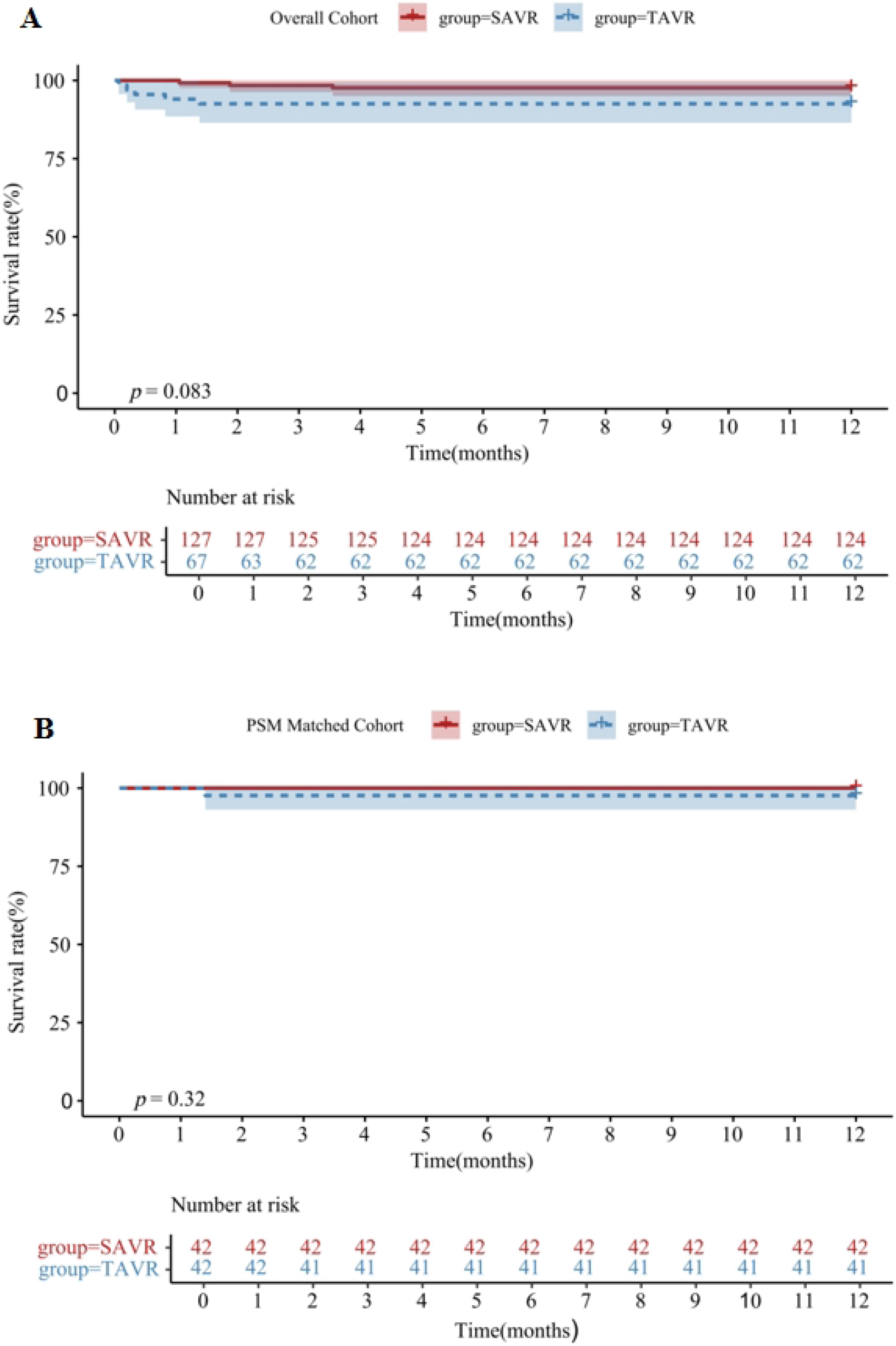
Kaplan-Meier curves for 1-year overall survival in patients with borderline pulmonary hypertension undergoing TAVR versus SAVR: (A) Overall Cohort analysis, (b) PSM Matched Cohort. Survival curves are obtained by using the KaplaneMeier method, and survival analysis is performed with the use of Cox proportional-hazard model. PSM, propensity score matching; TAVR, transcatheter aortic valve replacement; SAVR, surgical aortic valve replacement.

### Exploratory results

The crystalloid infusion volume in the TAVR group was greater than that in SAVR group (P<0.001). The intraoperative plasma infusion volume was greater in SAVR group (P=0.003), and there was more intraoperative allogeneic blood (P<0.001) and more blood loss (P<0.001). Compared with patients of SAVR group, participants of TAVR group had shorter operative duration (P<0.001), shorter hospital stay (P=0.001), earlier extubation time (P=0.006), and shorter intensive care unit stay (P<0.001). In addition, the hospitalization costs of patients of TAVR group were significantly higher than those of SAVR group (P<0.001). The above results were still consistent with the significant differences after PSM adjustment.

According to postoperative echocardiogram data, the incidence of mild perivalvular regurgitation in the TAVR group was 31 (46.2%), while in the SAVR group, it was 4 (3.1%) (P<0.001). After matching, 21 patients (50%) in TAVR group and 1 patient (2.4%) in SAVR group (P<0.001) were included. The incidence of mild regurgitation of the aortic valve orifice in TAVR was 41 (61.2%), while in SAVR group, it was 112 (88.2%) (P<0.001). After matching, 25 participants (59.5%) in TAVR and 40 participants (95.2%) in SAVR (P<0.001) were included. However, when the two types of aortic valve regurgitation data were integrated together, it was found that, no significant difference was found between the two groups. The differences about other exploratory results, containing the use of postoperative permanent pacemakers, between the TAVR and SAVR cohorts were not statistically significant and remained not significant after PSM adjustment (Table 5).

**Table 5.**
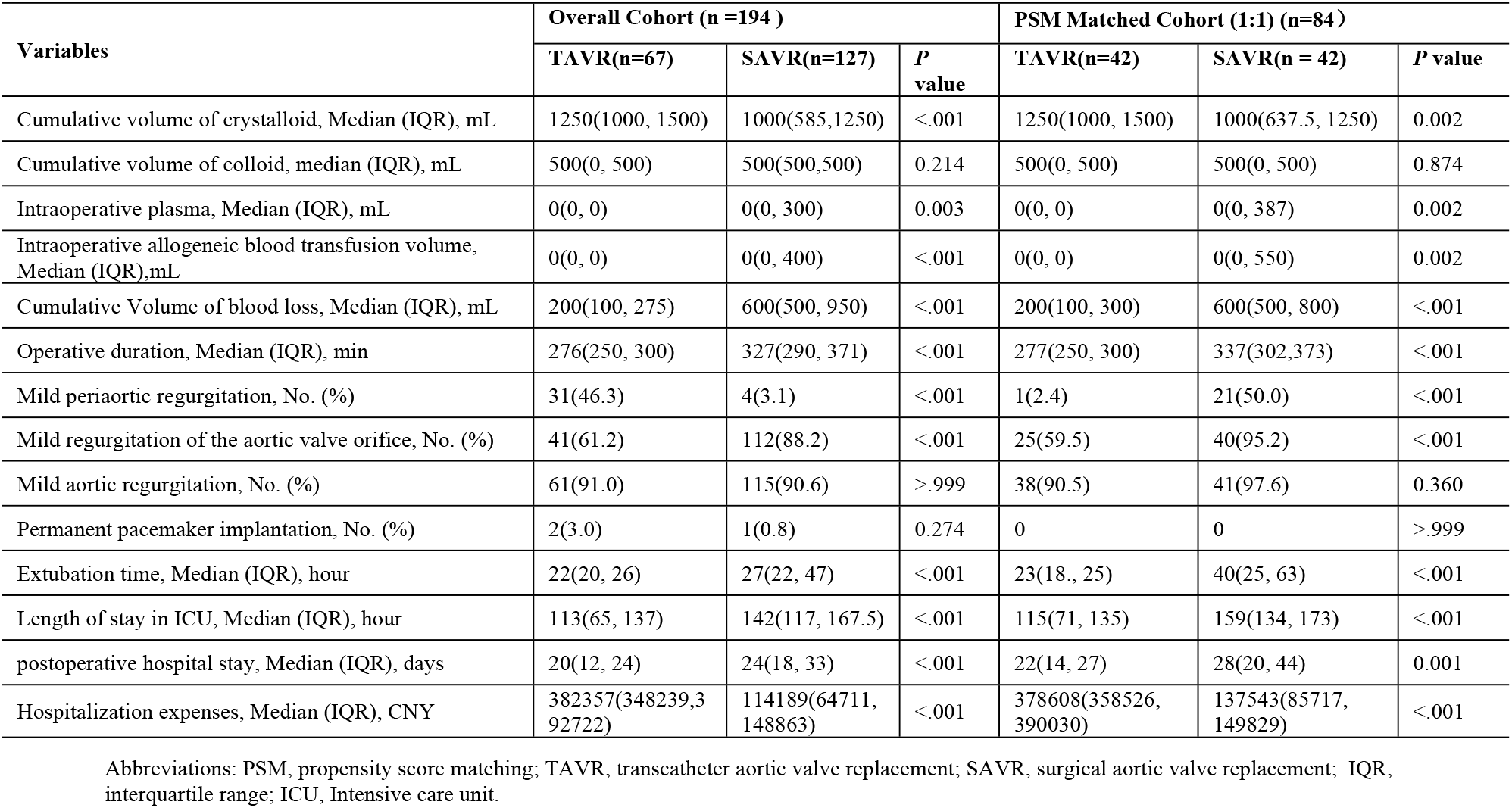
Exploratory results.

| Variables | Overall Cohort (n =194 ) |  |  | PSM Matched Cohort (1:1) (n=84) |  |  |
| --- | --- | --- | --- | --- | --- | --- |
|  | TAVR(n=67) | SAVR(n=127) | P value | TAVR(n=42) | SAVR(n = 42) | P value |
| Cumulative volume of crystalloid, Median (IQR), mL | 1250(1000, 1500) | 1000(585,1250) | <.001 | 1250(1000, 1500) | 1000(637.5, 1250) | 0.002 |
| Cumulative volume of colloid, median (IQR), mL | 500(0, 500) | 500(500,500) | 0.214 | 500(0, 500) | 500(0, 500) | 0.874 |
| Intraoperative plasma, Median (IQR), mL | 0(0, 0) | 0(0, 300) | 0.003 | 0(0, 0) | 0(0, 387) | 0.002 |
| Intraoperative allogeneic blood transfusion volume, Median (IQR),mL | 0(0, 0) | 0(0, 400) | <.001 | 0(0, 0) | 0(0, 550) | 0.002 |
| Cumulative Volume of blood loss, Median (IQR), mL | 200(100, 275) | 600(500, 950) | <.001 | 200(100, 300) | 600(500, 800) | <.001 |
| Operative duration, Median (IQR), min | 276(250, 300) | 327(290, 371) | <.001 | 277(250, 300) | 337(302,373) | <.001 |
| Mild periaortic regurgitation, No. (%) | 31(46.3) | 4(3.1) | <.001 | 1(2.4) | 21(50.0) | <.001 |
| Mild regurgitation of the aortic valve orifice, No. (%) | 41(61.2) | 112(88.2) | <.001 | 25(59.5) | 40(95.2) | <.001 |
| Mild aortic regurgitation, No. (%) | 61(91.0) | 115(90.6) | >.999 | 38(90.5) | 41(97.6) | 0.360 |
| Permanent pacemaker implantation, No. (%) | 2(3.0) | 1(0.8) | 0.274 | 0 | 0 | >.999 |
| Extubation time, Median (IQR), hour | 22(20, 26) | 27(22, 47) | <.001 | 23(18., 25) | 40(25, 63) | <.001 |
| Length of stay in ICU, Median (IQR), hour | 113(65, 137) | 142(117, 167.5) | <.001 | 115(71, 135) | 159(134, 173) | <.001 |
| postoperative hospital stay, Median (IQR), days | 20(12, 24) | 24(18, 33) | <.001 | 22(14, 27) | 28(20, 44) | 0.001 |
| Hospitalization expenses, Median (IQR), CNY | 382357(348239,392722) | 114189(64711, 148863) | <.001 | 378608(358526, 390030) | 137543(85717, 149829) | <.001 |

Abbreviations: PSM, propensity score matching; TAVR, transcatheter aortic valve replacement; SAVR, surgical aortic valve replacement; IQR, interquartile range; ICU, Intensive care unit.

## Discussion

Pulmonary hypertension, historically defined by a mean pulmonary arterial pressure ≥25 mmHg, is a well-established predictor of poor outcomes in patients undergoing TAVR for severe aortic stenosis [20, 21]. However, the recent proposal to lower the pulmonary hypertension diagnostic threshold to mean pulmonary arterial pressure of 20 mmHg. Once considered a clinically irrelevant variant of normal physiology, borderline pulmonary hypertension is now recognized as a transitional state preceding overt pulmonary hypertension, associated with subclinical right ventricular remodeling, diastolic dysfunction, and increased perioperative risk [9, 22]. In the context of aortic valve disease, even modest elevations in pulmonary pressure disrupt right ventricular, leaving patients vulnerable to hemodynamic instability during valve replacement, which is a gap that has long been overlooked in comparative effectiveness research on TAVR versus SAVR [9, 23].

This retrospective cohort study compared TAVR and SAVR-associated postoperative complications in patients with borderline pulmonary hypertension over 11 years. Multivariable analyses performed before and after matching demonstrated no significant differences between the two groups, implying that the selection of TAVR or SAVR does not impact 30-day primary composite complication rates or 1-year mortality in patients with borderline pulmonary hypertension. When individual complications were analyzed, even post-matching, the TAVR group showed a significantly greater incidence of cardiovascular complications than the SAVR group, but a significantly lower incidence of renal complications. Furthermore, the cumulative volume of blood loss and cumulative volume of fluid input, extubation time, intensive care unit stay and hospital stay in SAVR group were worse than those in TAVR group. Nevertheless, it is worth mentioning that the hospitalization expense of SAVR was significantly lower than that of TAVR.

Our results build on prior research by validating TAVR-SAVR equivalence on the incidence of 30-d composite complications and one-year mortality in a borderline pulmonary hypertension-specific cohort. Ahmad demonstrated comparable outcomes between TAVR and SAVR in mixed pulmonary hypertension populations, but their analysis did not stratify by borderline pulmonary hypertension [21]. Similarly, the UK TAVI Trial showed TAVR non-inferiority in older, higher-risk patients but excluded borderline pulmonary hypertension as a subgroup [23]. Our study fills this niche by showing that equivalence persists even in patients with the unique vulnerabilities of borderline pulmonary hypertension. Notably, our center’s practice of involving patients and families in treatment decision-making (consistent with real-world clinical workflows) enhances the external validity of our findings.

The higher incidence of cardiovascular complications in the TAVR group may be attributable to intraoperative hemodynamic perturbations, including rapid ventricular pacing and transient hypotension, which could precipitate right ventricular ischemia or acute dysfunction in patients with limited cardiac reserve. Conversely, the increased renal complication rate in the SAVR group likely reflects the systemic inflammatory response and microembolic burden associated with cardiopulmonary bypass, which may exacerbate subclinical cardiorenal syndrome, a common comorbidity in borderline pulmonary hypertension [9].

One notable finding requiring context is the pre-matching 30-day mortality difference (6.0% in TAVR vs. 0% in SAVR; P=0.013), which resolved after PSM. This pre-matching discrepancy likely reflects baseline differences in patient risk (e.g., older age or more comorbidities in the TAVR cohort, as seen in Table 1) rather than an inherent procedure risk. The post-matching absence of 30-day mortality and only 1 TAVR-related death at 1 year (vs. 0 in SAVR) aligns with long-term data showing no survival differences in low-to-intermediate risk patients receiving TAVR and SAVR [24, 25], including those with pulmonary hypertension. Jorgensen reported 8-year survival equivalence in low-risk aortic stenosis patients [24], and Çelik noted durable SAVR survival over 30 years [26]. These findings, combined with our data, suggest borderline pulmonary hypertension does not alter the long-term prognostic parity of the two procedures.

Beyond complication profiles, our results confirm previously reported procedural trade-offs: TAVR was associated with reduced operative time, less blood loss, earlier extubation, and shorter intensive care unit and hospital stays, while SAVR remained significantly less costly. These differences did not translate into divergent short-term clinical outcomes, supporting the expanded application of TAVR in patients with borderline pulmonary hypertension. However, the considerable financial burden of TAVR consumables remains a barrier preventing its widespread use, particularly in resource-limited settings [27].

These findings highlight the importance of integrating not only baseline pulmonary pressures but also right ventricular function, pulmonary vascular resistance, and renal reserve into preoperative risk stratification. Future prospective studies should investigate whether targeted perioperative interventions, such as right ventricular-protective ventilation strategies, selective pulmonary vasodilation, or renal preservation protocols, can mitigate procedure-specific complications in this high-risk population. Additionally, the long-term trajectory of pulmonary hemodynamics and right ventricular adaptation following TAVR versus SAVR in patients with borderline pulmonary hypertension remains poorly understood and warrants longitudinal investigation.

### Limitations

First, our study is retrospective, and it is difficult to avoid the possibility of data loss and incompleteness. Second, the sample size of the present study was relatively small, so additional prospective research involving larger patient cohorts is required to verify these findings in the subgroup of patients with borderline pulmonary hypertension. Additionally, since various transcatheter heart valves were included in this study, the prognosis may vary among different valves. Moreover, the inclusion of multiple valve types and the lack of stratification by aortic stenosis versus regurgitation may introduce heterogeneity. To validate our findings and enhance treatment algorithms, additional multicenter prospective trials with larger cohorts and standardized procedures are necessary.

## Conclusions

Our research suggested there was no significant discrepancy in 30-d primary composite complications or mortality, between patients with borderline pulmonary hypertension undertaking TAVR or SAVR. The TAVR group exhibited reduced intraoperative blood loss and blood transfusion requirements, earlier postoperative extubation, and shorter durations of stay in intensive care unit and hospital while SAVR group had significantly lower hospitalization expenses. Our findings support a personalized approach to valve replacement in this underrecognized but clinically important population.

## Data Availability

All relevant data are within the manuscript and its Supporting Information files.

## Supporting information

Table S1. Definitions of major postoperative complications

## Acknowledgments

We thank the participants from the staff of department of Anesthesiology to the First Affiliated Hospital of Soochow University and institute of Anesthesiology to Soochow University.

